# The Relationship between Cannabis Use and Cognition in People with Bipolar Disorder: A Systematic Scoping Review

**DOI:** 10.1101/2020.04.24.20078535

**Authors:** T. Jordan Walter, Nina Pocuca, Jared W. Young, Mark A. Geyer, Arpi Minassian, William Perry

**Affiliations:** Department of Psychiatry, University of California San Diego, 9500 Gilman Drive, La Jolla, CA 92093; Research Services, Veterans Administration San Diego HealthCare System, 3350 La Jolla Village Drive, San Diego CA, 92161; VA Center of Excellence for Stress and Mental Health, Veterans Administration San Diego HealthCare System, 3350 La Jolla Village Drive, San Diego CA, 92161

**Author notes:** Corresponding Author: Address: 410 Dickinson St, Office 6, San Diego CA, 92103.

## Abstract

Bipolar disorder (BD) and cannabis use are highly comorbid and are each associated with cognitive impairment. It is therefore important to understand the relationship between cannabis use and cognition in people with BD, as cannabis use in BD may be associated with greater cognitive impairment. We performed a scoping review to determine how much and what is currently known in this field. We systematically searched PubMed, Embase, CINAHL, Web of Science, and PsycINFO for studies on the relationship between cannabis and cognition in people with BD or relevant animal models. Six observational human studies met inclusion criteria. Two studies found cannabis use in BD was associated with better performance in some cognitive domains, while three studies found no association. One study found cannabis use in BD was associated with worse overall cognition. Overall, most identified studies suggest cannabis use is not associated with significant cognitive impairment in BD; however, the scope of knowledge in this field is limited, and more systematic studies are clearly required. Future studies should focus on longitudinal and experimental trials, as well as well-controlled observational studies with rigorous quantification of the onset, frequency, quantity, duration, and type of cannabis use.

## 1. INTRODUCTION

With increasing legalization and shifting attitudes regarding safety and acceptability, rates of cannabis use in the United States have risen in recent years (Hasin, 2018). Given these higher levels of use, understanding the effects of cannabis is becoming an increasingly important public health concern. Studying the effects of cannabis in psychiatric populations is especially relevant, as people with mental health conditions have higher rates of cannabis use than the general population (Lapham et al., 2017). People with bipolar disorder (BD) are notable for their high rates of cannabis use, with 71% of people with BD reporting a lifetime history of cannabis use compared to 27% of the general population (Agrawal et al., 2011). Furthermore, 17% of people with BD have a lifetime cannabis use disorder (CUD) compared to 6% of the general population, and 7.2% have a past-year CUD compared to 1.2% of the general population (Hasin et al., 2016; Hunt et al., 2016; Lev-Ran et al., 2013). Overall, these studies show that cannabis use is highly prevalent among people with BD. It is therefore important to understand the effects of cannabis in this population.

Previous studies have typically focused on the relationship between cannabis and mood in people with BD, finding that chronic cannabis use is associated with worse affective symptoms and clinical course (Baethge et al., 2008; de la Fuente-Tomás et al., 2020; Strakowski et al., 2007; Zorrilla et al., 2015); however, it is also important to understand the effects of cannabis on cognition in BD. Indeed, many people with BD exhibit cognitive deficits, including impairments in attention, executive function, fluency, learning, memory, and working memory (Bourne et al., 2013; Kurtz and Gerraty, 2009; Robinson et al., 2006; Young et al., 2019). These cognitive deficits can persist during euthymia (Robinson et al., 2006) and are a source of significant functional impairment independent of affective symptoms (Burdick et al., 2010). Importantly, cannabis use is itself associated with cognitive impairment (Broyd et al., 2016; Theunissen et al., 2015), and therefore has the potential to exacerbate the cognitive deficits of BD. Given the high rates of cannabis use among people with BD, understanding the effects of cannabis use in this population is important. Cannabis use in BD has the potential to worsen cognition, increase functional impairment, and exacerbate the significant societal costs associated with BD (Cloutier et al., 2018). The increasing availability of cannabis with legalization makes this matter timely and relevant. We therefore performed a scoping review to determine how much and what is currently known about the relationship between cannabis use and cognition in people with BD. We also sought to critically evaluate this literature, identify gaps in knowledge, and suggest directions for future research.

## 2. METHODS

### 2.1 Information sources and search strategy

We performed this systematic scoping review in accordance with the Preferred Reporting Items for Systematic Reviews and Meta-Analyses (PRISMA) guidelines (Liberati et al., 2009). Reports were identified by searching PubMed/MEDLINE, Embase, CINAHL, Web of Science, and PsycINFO for articles published through Oct 2019. We searched for reports on the relationship between cannabis and cognition in people with BD or relevant animal models with the help of a librarian experienced in systematic reviews. An example of the search strategy used in PubMed/MEDLINE is provided in Supplemental File 1.

### 2.2 Eligibility criteria and study selection

Eligible studies included those examining individuals 18 years or older with a bipolar spectrum disorder and a history of cannabis use who underwent cognitive assessment. Cognitive assessments of interest included, but were not limited to, attention, concentration, decision-making, executive function, general intelligence, learning, memory, processing speed, response inhibition, and working memory. We excluded studies with subjects with potentially confounding conditions such as neurological disorder, head/brain trauma, psychiatric illness other than BD, and active substance use disorders other than CUDs. We included studies that examined cannabinoids such as THC or cannabidiol (CBD) on cognition. Although our primary interest was comparing BD individuals with a cannabis use history to BD individuals without a cannabis use history, we were also interested in comparing these groups to cannabis users without BD and to healthy controls. To be as comprehensive as possible, we included all types of human studies examining the relationship between cannabis and cognition in BD. We also sought relevant animal studies. There were no restrictions by study setting or geographical location. We included only articles published in or translated into the English language.

### 2.3 Data collection process and data items

Database search results were uploaded to the internet-based EndNote software where duplicated results were deleted. All unique articles were then uploaded to Rayyan QCRI, an internet-based software that aids with systematic reviews. A level 1 assessment was performed in which two reviewers, TJW and NP, independently read the titles and abstracts of each report. They came to a consensus regarding which reports were eligible for further review based on the predetermined inclusion and exclusion criteria described above. Next, a level 2 assessment was performed in which TJW and NP independently read the full text reports and determined whether they met the inclusion and exclusion criteria. They came to a consensus regarding which reports would be included in data abstraction and risk of bias assessment. TJW and NP independently extracted data from eligible reports using standardized forms. Extracted information included the first author, publication year, diagnoses of the participants, mood status at time of study, medication status at time of study, cannabinoid use/exposure, number of subjects (n), cognitive assessments performed, and main findings. TJW and NP compared data extraction and came to a consensus. Disagreements were resolved by discussion.

### 2.4 Risk of bias in individual studies

The risk of bias of individual studies was assessed using the Agency for Healthcare Research and Quality Evidence-based Practice Center Methods Guide (Viswanathan et al., 2012). Studies were rated on selection bias, performance bias, attrition bias, detection bias, and reporting bias as applicable. These ratings were carried out independently by TJW and NP using a standardized form. Consensus regarding risk of bias assessments was reached through discussion. Disagreements were resolved by discussion.

## 3. RESULTS

### 3.1 Search results

The initial search results yielded 827 reports (Fig 1). Once duplicates were removed, 668 unique articles remained. The majority of reports did not meet the basic inclusion criteria of studying the relationship between cannabis and cognition in BD. Most articles addressed one or two, but not all three of the main concepts of cannabis, cognition, and BD. For example, many articles were excluded for examining the relationship between cannabis and cognition in people with schizophrenia rather than people with BD. Of the 668 unique articles, 18 (2.7%) were considered relevant based on a Level 1 title and abstract screening. Of these, six articles (0.9%) were selected as meeting eligibility criteria based on a Level 2 full text screening (Fig 1). The most common reasons for excluding articles at level 2 included the report being a conference abstract that duplicated a published study and the report not specifically assessing cognition.

**Figure 1.**
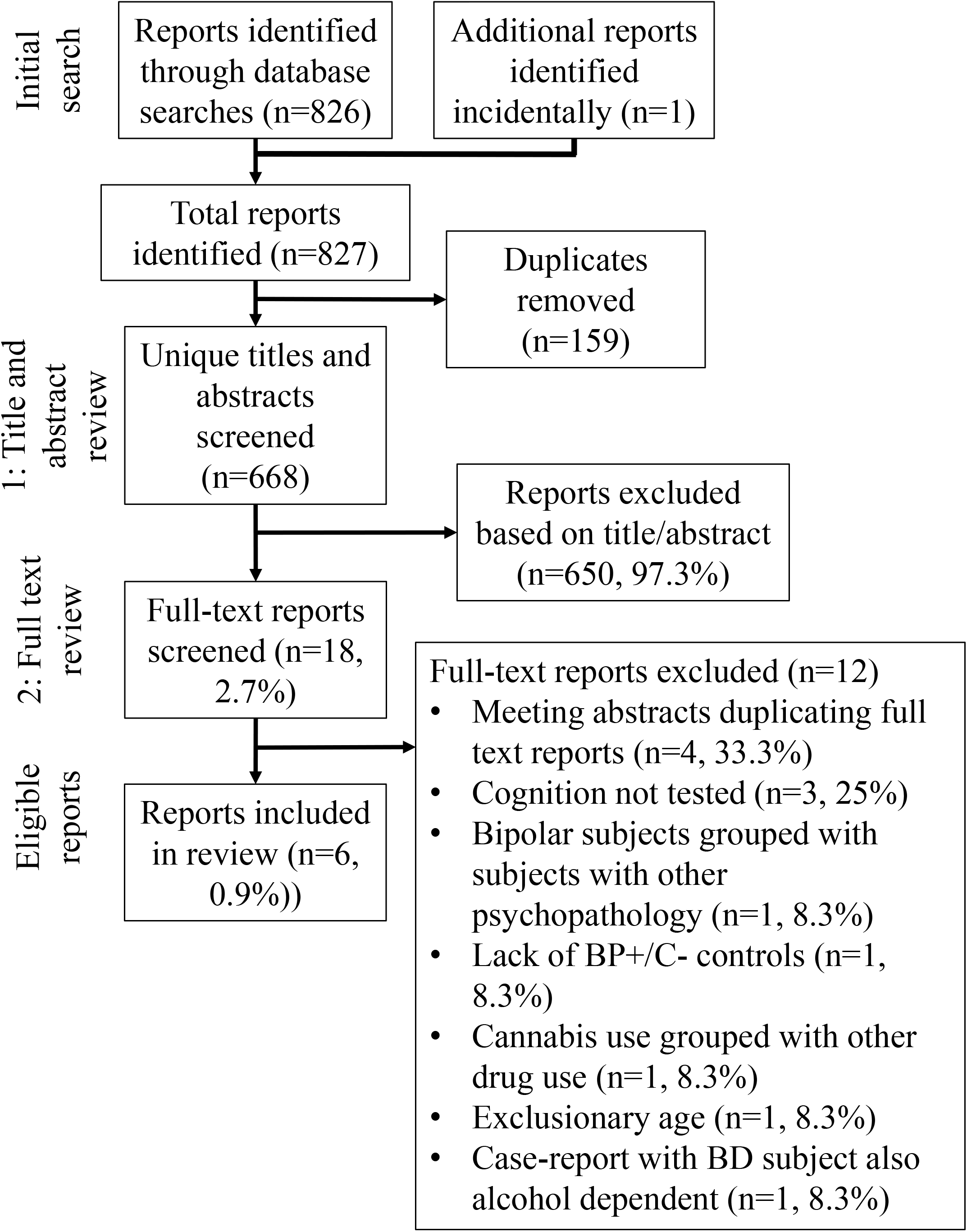
PRISMA Flowchart of Study Selection.

### 3.2 Human studies

We found six observational reports that studied the association between cannabis use and cognition in people with BD (Table 1). Two of these studies concluded that a history of cannabis use in people with BD was associated with better performance in certain cognitive domains. For example, Braga et al. examined the association between a CUD history and cognition in people with BD. This study found that a history of CUD in BD was associated with better attention, working memory, and processing speed compared to people with BD with no CUD history (Table 1). There was no association between cannabis use and general intelligence, verbal fluency, or learning and memory. In a different study, Ringen et al. compared people with BD who used cannabis in the past six months (meeting criteria for a CUD was not required in this study) to people with BD who did not use cannabis. Results showed that cannabis use in the past six months was associated with better general intelligence and verbal fluency in people with BD (Table 1). Unlike Braga et al., Ringen et al. did not find that cannabis-using people with BD performed better on attention, processing speed, or working memory measures compared to non-cannabis using people with BD. Overall, these two studies suggest a history of cannabis use in BD may be associated with better performance in certain cognitive domains, although the results of the studies differed with respect to the specific cognitive domains affected.

Three studies concluded there was no association between cannabis use history and cognition in people with BD. One of these studies focused on adolescent cannabis use and cognition in adulthood (Abush et al., 2018). This study found that adolescent cannabis use was not associated with different cognitive performance in people with BD during adulthood. Cognitive domains assessed included general intelligence, learning, memory, attention, working memory, motor speed, verbal fluency, processing speed, and executive function (Table 1). A second study examined people with BD with chronic cannabis (Sagar et al., 2016). In this study, the cannabis-using people with BD performed no differently than non-cannabis using people with BD in general intelligence, executive function, attention, processing speed, working memory, verbal fluency, learning, memory, and visuoperception (Table 1). Finally, we also identified an abstract reporting that individuals with Bipolar I Disorder (BD I), Bipolar II Disorder (BD II), or BD Not Otherwise Specified (NOS) with premorbid or current cannabis use had similar cognitive performance to non-cannabis using people with BD (Lagerberg et al., 2019) (Table 1). Overall, these reports indicated that a history of cannabis was not associated with cognitive changes in people with BD.

Finally, there was one study reporting that cannabis use was associated with worse performance in multiple cognitive domains in people with BD (Halder at al. 2016). This study was unique among the six, as it was the only study to examine solely acutely symptomatic participants. Results showed that symptomatic participants with BD and current cannabis dependence had worse verbal fluency, attention, processing speed, working memory, executive function and global cognition than symptomatic, non-cannabis-dependent people with BD (Table 1). Therefore, cannabis use in people with acutely symptomatic BD may worsen performance in certain cognitive domains.

### 3.3 Animal studies

No animal studies were found that examined the relationship between cannabis or cannabinoids and cognition in animal models relevant to BD; however, we identified two studies investigating the effects of cannabinoids on mania-like behavior, specifically locomotor hyperactivity. One study examined the Gria1-/-mouse strain that lacks the GluA1 AMPA receptor subunit. These mice exhibited mania-like hyperactivity when placed in a novel environment. This hyperactivity was blunted by both systemic and intra-hippocampal administration of CBD (Aitta-aho et al., 2019). Another study, however, failed to find any effect of CBD on locomotor activity in a different model of bipolar mania. This model used sub-chronic D-amphetamine (AMPH) to induce mania-like hyperactivity in rats. CBD failed to prevent or reverse AMPH-induced locomotor activity, despite normalizing AMPH-induced molecular markers in the brain (Valvassori et al., 2011). Overall, the scarcity of animal studies examining the relationship between cannabis or cannabinoids and cognition in animal models relevant to BD highlights the need for further animal research.

### 3.4 Risk of bias in individual studies

Studies were rated on selection bias, performance bias, attrition bias, detection bias, and reporting bias as applicable. A thorough review of the bias in these individual studies can be found in the Supplementary File 2. The studies varied in their selection and performance bias, with Braga et al. and Ringen et al. having a low risk of these biases. There was concern Halder et al. may have a high risk of selection bias because they discussed neither the percentage of men and women in their groups, nor the medication status of their groups. It was therefore unclear whether differences in these factors could have contributed to their positive results. The risk of selection and performance bias was unable to be determined in the other studies because it was unclear whether the analyses controlled for important confounding and modifying variables or whether the strategies for recruiting participants into the study differed across study groups. Attrition bias was not a concern across these studies given their cross-sectional nature. Detection bias was a concern across all studies due to failure to discuss blinding. Finally, reporting bias was unable to be fully determined due to lack of registered protocols, although all studies examined the endpoints described in their methods.

## 4. DISCUSSION

### 4.1 Human studies

We identified six human studies. Results of these studies varied, potentially as a result of multiple factors. Firstly, studies differed on the subtypes of BD they included. There was no clear association between the subtypes of BD examined and the different outcomes between the studies. Secondly, studies differed on the mood status of the participants included. These studies mostly examined stable patients, with one study including some patients who were symptomatic (Braga et al., 2012), and one study examining only acutely symptomatic participants (Halder et al., 2016). Halder et al. was also the only study to find that cannabis use was associated with worse cognition in people with BD. It is possible cannabis use interacted with acutely symptomatic BD to worsen cognition, an effect not seen in stable patients. Thirdly, some studies did not report the number or types of medications used. No study examined the relationship between medication, cannabis use, and cognition in BD, making it unclear whether differences in medication status contributed to the different results of the studies. Finally, it is worth noting these studies included both men and women, although most did not examine sex differences. Only Braga et al. examined sex differences and found no effect of sex on any of the neurocognitive measures. Lack of understanding of sex differences is an important gap in our understanding, as previous studies find evidence that cannabis has different effects in people with BD depending on sex (de la Fuente-Tomás et al., 2020).

Perhaps the most striking difference between the studies is their different criteria for cannabis use. Average duration, frequency, and quantity of cannabis use were not specified for most of the studies. It is possible differences in these parameters may be associated with discrepant effects on cognition. Furthermore and importantly, many studies allowed for varying abstinence periods between last cannabis use and cognitive testing that may have contributed to contrasting outcomes. Abstinence from cannabis use is associated with normalized cognition in healthy controls (Schreiner and Dunn, 2012; Scott et al., 2018). Therefore, prolonged abstinence in these BD studies may have allowed for normalized cognition. Also, because many studies did not verify the last time cannabis was used before cognitive testing, it is possible that withdrawal or even intoxication effects contributed to the variation in results. The type and potency of the cannabis used is also not reported. The primary cannabinoids found in cannabis, such as THC and CBD, are known to have different effects on cognition (Morgan et al., 2018), therefore, use of different cannabis strains with varying levels of cannabinoids may have diverse effects on cognition in BD. Overall, the marked methodological differences between these studies likely contributed to the varying results.

### 4.2 Animal studies

There were no reports examining the relationship between cannabis or cannabinoids and cognition in animal models relevant to BD, showing that more work in this area is needed. Previous studies have used animal models of bipolar mania, such as dopamine transporter (DAT) knockdown mice, to assess cognition (van Enkhuizen et al., 2014; Young et al., 2019). Other animal models may also be useful in studying the relationship between cannabis and cognition in BD, such as the ClockΔ19 mouse (Kristensen et al., 2018) or the amphetamine-sensitization model (Fries et al., 2015). Experiments in relevant animal models would enable testing of mechanistic hypotheses that could not be done in humans, as well as testing of novel therapeutics to treat the cognitive deficits of BD.

### 4.3 Risk of bias assessment

Studies varied in their levels of bias. Braga et al. and Ringen et al. were relatively low in overall bias, suggesting we can view their results with a higher level of confidence. Interestingly, both of these studies found that cannabis improved cognition in BD. Halder et al. had a higher risk of bias due to concerns regarding selection bias. The sex and medication status of the groups in this study were not reported, leaving it unclear whether these factors could have contributed to the positive findings. Notably, this was the only study finding cannabis worsened cognition in BD. All other studies had an intermediate overall risk of bias and found no effect of cannabis on cognition in BD. Therefore, our risk of bias assessment indicates that the literature currently supports cannabis not having a markedly detrimental effect on cognition in BD.

### 4.4 Future research

Our systematic review suggests many areas for future research. All of the studies identified were cross-sectional studies. An experimental study would provide insight into causality regarding the effects of cannabis on cognition in BD. Future studies should also consider the effects of cannabis or cannabinoids on neurotransmitter systems involved in cognition in people with BD, such as the acetylcholine or dopamine system. Such a direction would provide insight into the underlying neurobiology of cannabis and cognition in BD. Another area open for research is a well-controlled observational study with more rigorous quantification of the onset, frequency, quantity, duration, and type of cannabis used, as well as the reasons for use. It was also unclear from the identified studies how much time elapsed between last cannabis use and cognitive testing. Therefore, a study that defines a standardized time post-use for cognitive testing that avoided both intoxication and withdrawal effects is important. Studying the effects of individual cannabinoids such as THC and CBD on cognition in BD would provide more detailed insights into the effects of cannabis in this population. Finally, the study by Halder et al. suggests cannabis use may have different effects on cognition in BD depending on the affective state. Therefore, another interesting study could compare the effects of cannabis on cognition in states of mania and depression.

### 4.5 Limitations

The limitations of this review include the number of studies that met inclusion and exclusion criteria. Only six studies were identified, which weakens the conclusions that can be drawn regarding the association between cannabis use and cognition in BD. Also, given that each of the identified studies was observational, it is impossible to draw conclusions regarding causality. Finally, as discussed above, the significant heterogeneity of definitions of cannabis use among these studies makes it difficult to draw clear conclusions.

### 4.6 Conclusion

This systematic scoping review sought to determine how much and what is known about the relationship between cannabis and cognition in people with BD. We identified a small number of studies, most of which suggest a history of cannabis use was not associated with significantly worse cognition in BD. However, given the relative lack of studies in this field, the clinical implications of this research currently remain unclear. The death of knowledge in this areas highlights the clear need for further research. Such studies should focus on longitudinal designs or experimental designs that will enable conclusions to be drawn regarding causality. Well-controlled observational studies with rigorous quantification of the onset, duration, amount, type, and frequency of cannabis used, as well as the reasons for use are also essential. Given the extensive cannabis use among individuals with BD and the increasing availability of cannabis, studying the relationship between cannabis and cognition in BD is becoming increasingly important. There are many gaps in our knowledge of this area, with ample opportunities for future research.

## Data Availability

Data will be made available with the manuscript.

## ACKNOWLEDGEMENTS

We would like to acknowledge Karen Heskett for her assistance with the systematic review.

## AUTHOR CONTRIBUTIONS

Conceptualization: WP, AM, and TJW; Data curation: TJW and NP; Formal Analysis: TJW and NP; Funding Acquisition: WP, AM, and JWY; Investigation: TJW and NP; Writing – original draft: TJW; Writing – review and editing: TJW, NP, JWY, MAG, AM, and WP.

## SUPPLEMENTARY MATERIAL

File 1. Detailed Risk of Bias Assessment for Selected Studies

